## Supplementary material for "Combinations of genes at the 16p11.2 and 22q11.2 CNVs contribute to neurobehavioral traits": Note_S1.docx

Principal investigators of the Psychiatric Genomics Consortium

**Autism Working Group**

Aravinda Chakravarti

Center for Human Genetics and Genomics, New York University School of Medicine, New York, New York, USA

Mark J, Daly

Stanley Center for Psychiatric Research, The Broad Institute of Harvard and M.I.T, Cambridge, MA, USA

Analytic and Translational Genetics Unit, Massachusetts General Hospital, Boston, USA

Institute for Molecular Medicine Finland, University of Helsinki, Helsinki, Finland

Daniel H, Geschwind

Department of Neurology and Center for Neurobehavioral Genetics, University of California Los Angeles, Los Angeles, CA

Joachim Franz Hallmayer

Department of Psychiatry and Behavioral Sciences, Stanford University School of Medicine, Stanford, CA

Aarno Palotie

Psychiatric and Neurodevelopmental Genetics Unit, Massachusetts General Hospital, Boston, MA, USA
Institute for Molecular Medicine Finland, FIMM, Helsinki, Finland
The Broad Institute of MIT and Harvard, Cambridge, MA, USA

Guy A. Rouleau

Department of Human Genetics, McGill University, Montreal, QC, Canada

Department of Neurology and Neurosurgery, Montreal Neurological Institute, McGill University, Montreal, QC, Canada

John E. Spiro

Simons Foundation, New York, New York

Joseph D. Buxbaum

Department of Human Genetics, Icahn School of Medicine at Mount Sinai, New York, NY, USA
Department of Psychiatry, Icahn School of Medicine at Mount Sinai, New York, NY, USA.
Friedman Brain Institute, Icahn School of Medicine at Mount Sinai, New York, NY, USA
Department of Neuroscience, Icahn School of Medicine at Mount Sinai, New York, NY, USA

Lauren A. Weiss

Department of Psychiatry, University of California San Francisco, San Francisco, CA, USA

Institute for Human Genetics, University of California San Francisco, San Francisco, CA, USA

Weill Institute for Neurosciences, University of California San Francisco, San Francisco, CA, USA

**Bipolar Working Group**

Rolf Adolfsson

Department of Clinical Sciences, Psychiatry, Umeå University Medical Faculty, Umeå, SE

Ole A. Andreassen

Div Mental Health and Addiction, Oslo University Hospital, Oslo, NO

NORMENT, University of Oslo, Oslo, NO

Martin Alda, Gustavo Turecki, Guy A. Rouleau

Department of Psychiatry, Dalhousie University, Halifax, NS, CA;
National Institute of Mental Health, Klecany, CZ

Nicholas James Bass, Andrew McQuillin

Division of Psychiatry, University College London, London, GB

Joanna M. Biernacka, Mark Frye

Department of Health Sciences Research, Mayo Clinic, Rochester, MN, US
Department of Psychiatry & Psychology, Mayo Clinic, Rochester, MN, US

Douglas H. R. Blackwood

Division of Psychiatry, University of Edinburgh, Edinburgh, GB

Michael Boehnke, Laura J. Scott

Center for Statistical Genetics and Department of Biostatistics, University of Michigan, Ann Arbor, MI, US

Sven Cichon

Department of Biomedicine, University of Basel, Basel, Switzerland

Institute of Human Genetics, University of Bonn, School of Medicine & University Hospital Bonn, Bonn, Germany

Institute of Medical Genetics and Pathology, University Hospital Basel, Basel, Switzerland

Institute of Neuroscience and Medicine (INM-1), Research Centre Jülich, Jülich, Germany

Ashley L. Comes

Institute of Psychiatric Phenomics and Genomics (IPPG), University Hospital, LMU Munich, Munich, Germany

International Max Planck Research School for Translational Psychiatry (IMPRS-TP), Munich, Germany

Nicholas Craddock, Arianna Di Florio, Ian Jones

Medical Research Council Centre for Neuropsychiatric Genetics and Genomics, Division of Psychological Medicine and Clinical Neurosciences, Cardiff University, Cardiff, GB

Aiden Corvin

Neuropsychiatric Genetics Research Group, Dept of Psychiatry and Trinity Translational Medicine Institute, Trinity College Dublin, Dublin, IE

Franziska Degenhardt

Department of Child and Adolescent Psychiatry, Psychosomatics and Psychotherapy, University Hospital Essen, University of Duisburg-Essen, Duisburg, Germany

Institute of Human Genetics, University of Bonn, School of Medicine & University Hospital Bonn, Bonn, Germany

Andreas J. Forstner

Centre for Human Genetics, University of Marburg, Marburg, Germany

Institute of Human Genetics, University of Bonn, School of Medicine & University Hospital Bonn, Bonn, Germany

Janice M. Fullerton, Peter R. Schofield

Neuroscience Research Australia, Sydney, NSW, AU

School of Medical Sciences, University of New South Wales, Sydney, NSW, AU

Maria Grigoroiu-Serbanescu

Biometric Psychiatric Genetics Research Unit, Alexandru Obregia Clinical Psychiatric Hospital, Bucharest, Romania

José Guzman-Parra

Mental Health Department, University Regional Hospital, Biomedicine Institute (IBIMA), Málaga, Spain

Joanna Hauser

Department of Psychiatry, Laboratory of Psychiatric Genetics, Poznan University of Medical Sciences, Poznan, Poland

Lisa Jones

Department of Psychological Medicine, University of Worcester, Worcester, GB

John Kelsoe

Department of Psychiatry, University of California San Diego, La Jolla, CA, US

George Kirov

Medical Research Council Centre for Neuropsychiatric Genetics and Genomics, Division of Psychological Medicine and Clinical Neurosciences, Cardiff University, Cardiff, GB

Manolis Kogevinas

ISGlobal, Barcelona, Spain

Mikael Landén

Department of Medical Epidemiology and Biostatistics, Karolinska Institutet, Stockholm, SE

Institute of Neuroscience and Physiology, University of Gothenburg, Gothenburg, SE

Marion Leboyer

Faculté de Médecine, Université Paris Est, Créteil, FR
Department of Psychiatry and Addiction Medicine, Assistance Publique - Hôpitaux de Paris, Paris, FR
INSERM, Paris, FR

Jolanta Lissowska

Cancer Epidemiology and Prevention, M. Sklodowska-Curie National Research Institute of Oncology, Warsaw, Poland

Nicholas G. Martin

Genetics and Computational Biology, QIMR Berghofer Medical Research Institute, Brisbane, QLD, AU

School of Psychology, The University of Queensland, Brisbane, QLD, AU

Fermin Mayoral

Mental Health Department, University Regional Hospital, Biomedicine Institute (IBIMA), Málaga, Spain

Richard M. Myers

HudsonAlpha Institute for Biotechnology, Huntsville, AL, US

Philip B. Mitchell

Neuroscience Research Australia, Sydney, NSW, AU

Bertram Müller-Myhsok

Department of Translational Research in Psychiatry, Max Planck Institute of Psychiatry, Munich, Germany

Munich Cluster for Systems Neurology (SyNergy), Munich, Germany

University of Liverpool, Liverpool, United Kingdom

Markus M. Nöthen

Institute of Human Genetics, University of Bonn, School of Medicine & University Hospital Bonn, Bonn, Germany

Carlos N. Pato

Institute for Genomic Health, SUNY Downstate Medical Center College of Medicine, Brooklyn, NY, US
College of Medicine Institute for Genomic Health, SUNY Downstate Medical Center College of Medicine, Brooklyn, NY, US

Andreas Reif

Department of Psychiatry, Psychosomatic Medicine and Psychotherapy, University Hospital Frankfurt, Frankfurt am Main, Germany

Marcella Rietschel

Department of Genetic Epidemiology in Psychiatry, Central Institute of Mental Health, Medical Faculty Mannheim, Heidelberg University, Mannheim, DE

Sabrina Schaupp

Institute of Psychiatric Phenomics and Genomics (IPPG), University Hospital, LMU Munich, Munich, Germany

Lea Sirignano

Department of Genetic Epidemiology in Psychiatry, Central Institute of Mental Health, Medical Faculty Mannheim, Heidelberg University, Mannheim, Germany

Thomas G. Schulze

Institute of Psychiatric Phenomics and Genomics (IPPG), University Hospital, LMU Munich, Munich, Germany

Department of Psychiatry and Behavioral Sciences, Johns Hopkins University School of Medicine, Baltimore, MD, USA

Department of Genetic Epidemiology in Psychiatry, Central Institute of Mental Health, Medical Faculty Mannheim, Heidelberg University, Mannheim, Germany

Department of Psychiatry and Psychotherapy, University Medical Center Göttingen, Göttingen, Germany

Department of Psychiatry and Behaioral Sciences, SUNY Upstate Medical University, Syracuse, NY, USA

Jordan W. Smoller

Stanley Center for Psychiatric Research, Broad Institute, Cambridge, MA, US;
Department of Psychiatry, Massachusetts General Hospital, Boston, MA, US;
Psychiatric and Neurodevelopmental Genetics Unit (PNGU), Massachusetts General Hospital, Boston, MA, US

Fabian Streit

Department of Genetic Epidemiology in Psychiatry, Central Institute of Mental Health, Medical Faculty Mannheim, Heidelberg University, Mannheim, Germany

Patrick F. Sullivan

Department of Medical Epidemiology and Biostatistics, Karolinska Institutet, Stockholm, SE

Department of Genetics, University of North Carolina at Chapel Hill, Chapel Hill, NC, US

Department of Psychiatry, University of North Carolina at Chapel Hill, Chapel Hill, NC, US

Beata Świątkowska

Department of Environmental Epidemiology, Nofer Institute of Occupational Medicine, Lodz, Poland

**Schizophrenia Working Group**

Rolf Adolfsson

Department of Clinical Sciences, Psychiatry, Umeå University Medical Faculty, Umeå, SE

Ole A. Andreassen

Div Mental Health and Addiction, Oslo University Hospital, Oslo, NO;
NORMENT, University of Oslo, Oslo, NO

Celso Arango

Centro de Investigación Biomédica en Red en Salud Mental (CIBERSAM), Madrid, Spain.

Department of Child and Adolescent Psychiatry, Institute of Psychiatry and Mental Health, General Universitario Gregorio Marañón, School of Medicine, Universidad Complutense, IiSGM, Madrid, Spain.

E. Cem Atbaşoğlu, Meram Can Saka

Ankara University School of Medicine

Bernhard Baune

University Hospital of Psychiatry and Psychotherapy, University of Münster, Germany

Martin Begemann, Hannelore Ehrenreich, Agnes A. Steixner-Kuma

Clinical Neuroscience, Max Planck Institute of Experimental Medicine, Göttingen, Germany

Sintia Belangero‬

Laboratory of Integrative Neuroscience, Universidade Federal de São Paulo, São Paulo, Brazil

Department of Morphology and Genetics, Laboratorio de Genetica, Universidade Federal de São Paulo, São Paulo, Brazil

Douglas H. R. Blackwood

Division of Psychiatry, University of Edinburgh, Edinburgh, GB

Anders D. Børglum

The Lundbeck Foundation Initiative for Integrative Psychiatric Research, iPSYCH, Denmark
Centre for Integrative Sequencing, iSEQ, Aarhus University, Aarhus, Denmark
Department of Biomedicine, Aarhus University, Aarhus, Denmark
Department P, Aarhus University Hospital, Risskov, Denmark.

David Braff

University of California, San Diego, School of Medicine

Elvira Bramon

University College London, UK

Joseph D. Buxbaum

Department of Human Genetics, Icahn School of Medicine at Mount Sinai, New York, NY, USA
Department of Psychiatry, Icahn School of Medicine at Mount Sinai, New York, NY, USA.
Friedman Brain Institute, Icahn School of Medicine at Mount Sinai, New York, NY, USA
Department of Neuroscience, Icahn School of Medicine at Mount Sinai, New York, NY, USA

Dominique Campion, Claudine Laurent-Levinson

Groupe de Recherche sur la Schizophrénie, UER Médecine Rouen, Rouen, France

Jorge A Cervilla

Department of Psychiatry, San Cecilio University Hospital, University of Granada, Granada, Spain.

Aiden Corvin

Neuropsychiatric Genetics Research Group, Dept of Psychiatry and Trinity Translational Medicine Institute, Trinity College Dublin, Dublin, IE

Ariel Darvasi

Department of Genetics, The Hebrew University of Jerusalem, Jerusalem, Israel

Marta Di Forti

King’s College London

Enrico Domenici

Department of Cellular, Computational and Integrative Biology, University of Trento, Trento, Italy

Hannelore Ehrenreich

Max Planck Institute for Experimental Medicine

Tõnu Esko

Medical and Population Genetics Program, Broad Institute of MIT and Harvard, Cambridge, MA, USA.
Division of Endocrinology and Center for Basic and Translational Obesity Research, Boston Children's Hospital, Boston, MA, USA
Department of Genetics, Harvard Medical School, Boston, MA, USA
Estonian Genome Center, University of Tartu, Tartu, Estonia

Anna Gareeva, Elza Khusnutdinova

Institute of Biochemistry and Genetics, Ufa Scientific Center of Russian Academy of Sciences

Micha Gawlik

University of Wuerzburg

Pablo V. Gejman

Department of Psychiatry and Behavioral Neuroscience, University of Chicago, Chicago, IL, USA
Department of Psychiatry and Behavioral Sciences, NorthShore University HealthSystem, Evanston, IL, USA

Ina Giegling, Dan Rujescu

Department of Psychiatry, University of Halle, Halle, Germany

Vera V Golimbet

Department of Clinical Genetics, Mental Health Research Center

Christina M. Hultman

Department of Medical Epidemiology and Biostatistics, Karolinska Institutet, Stockholm,Sweden

Nakao Iwata

Department of Psychiatry, Fujita Health University School of Medicine, Toyoake, Aichi, Japan

Erik G. Jönsson

Department of Clinical Neuroscience, Karolinska Institutet, Stockholm, Sweden

George Kirov

Medical Research Council Centre for Neuropsychiatric Genetics and Genomics, Division of Psychological Medicine and Clinical Neurosciences, Cardiff University, Cardiff, GB

Marie-Odile Krebs

Service de Psychiatrie, Centre Hospitalier Sainte-Anne

Todd Lencz, Ariel Darvasi

The Feinstein Institute for Medical Research, Manhasset, NY, USA

The Hofstra NS-LIJ School of Medicine, Hempstead, NY, USA

The Zucker Hillside Hospital, Glen Oaks, NY, USA

Douglas F. Levinson

Department of Psychiatry and Behavioral Sciences, Stanford University, Stanford, CA, USA.

Jianjun Liu

Human Genetics, Genome Institute of Singapore, A*STAR, Singapore
Saw Swee Hock School of Public Health, National University of Singapore, Singapore

Anil K. Malhotra

The Feinstein Institute for Medical Research, Manhasset, NY, USA
The Hofstra NS-LIJ School of Medicine, Hempstead, NY, USA
The Zucker Hillside Hospital, Glen Oaks, NY, USA

Andrew McIntosh

University of Edinburgh, Royal Edinburgh Hospital

Andrew McQuillin

University College London

Paulo R. Menezes, Sintia Belangero‬

Departamento de Medicina Social, Faculdade de Medicina da Universidade de São Paulo

Bryan J .Mowry

Queensland Brain Institute, The University of Queensland, Brisbane, Queensland, Australia
Queensland Centre for Mental Health Research, University of Queensland, Brisbane, Queensland, Australia

Markus M. Nöthen

Institute of Human Genetics, University of Bonn, School of Medicine & University Hospital Bonn, Bonn, Germany

Vishwajit L Nimgaonkar

Department of Psychiatry, University of Pittsburgh

Michael C. O'Donovan, Michael J. Owen, James T.R. Walters

MRC Centre for Neuropsychiatric Genetics and Genomics, Institute of Psychological Medicine and Clinical Neurosciences, School of Medicine, Cardiff University, Cardiff, UK.

National Centre for Mental Health, Cardiff University, Cardiff, Wales

Roel A. Ophoff

Psychiatry, UMC Utrecht Brain Center Rudolf Magnus, Utrecht, NL
Human Genetics, University of California Los Angeles, Los Angeles, CA, US
Center for Neurobehavioral Genetics, University of California Los Angeles, Los Angeles, CA, US

Aarno Palotie

Psychiatric and Neurodevelopmental Genetics Unit, Massachusetts General Hospital, Boston, MA, USA
Institute for Molecular Medicine Finland, FIMM, Helsinki, Finland
The Broad Institute of MIT and Harvard, Cambridge, MA, USA

Carlos N. Pato

Institute for Genomic Health, SUNY Downstate Medical Center College of Medicine, Brooklyn, NY, US
College of Medicine Institute for Genomic Health, SUNY Downstate Medical Center College of Medicine, Brooklyn, NY, US

Tracey L. Petryshen

Department of Psychiatry, Harvard Medical School, Boston, MA, USA
The Broad Institute of MIT and Harvard, Cambridge, MA, USA
Center for Human Genetic Research and Department of Psychiatry, Massachusetts General Hospital, Boston, MA, USA

Marcella Rietschel

Department of Genetic Epidemiology in Psychiatry, Central Institute of Mental Health, Medical Faculty Mannheim, Heidelberg University, Mannheim, DE

Brien P. Riley

Virginia Institute for Psychiatric and Behavioral Genetics, Departments of Psychiatry and Human and Molecular Genetics, Virginia Commonwealth University, Richmond, VA, USA

Stephan Ripke

Analytic and Translational Genetics Unit, Massachusetts General Hospital, Boston, MA, USA.

Stanley Center for Psychiatric Research, Broad Institute of MIT and Harvard, Cambridge, MA, USA.

Dan Rujescu

Ludwig-Maximilians-University Munich

Alessandro Serretti

University of Bologna

Pak C. Sham

Centre for Genomic Sciences, State Ket Laboratory for Brain and Cognitive Sciences, and Department of Psychiatry, Li Ka Shing Faculty of Medicine, The University of Hong Kong, Hong Kong SAR, PR China

David St Clair

University of Aberdeen, Institute of Medical Sciences, Aberdeen, Scotland, UK.

Patrick F. Sullivan

Department of Medical Epidemiology and Biostatistics, Karolinska Institutet, Stockholm, SE

Department of Genetics, University of North Carolina at Chapel Hill, Chapel Hill, NC, US

Department of Psychiatry, University of North Carolina at Chapel Hill, Chapel Hill, NC, US

Jim van Os

Psychiatric Epidemiology and Public Mental Health at Utrecht University Medical Centre, the Netherlands

James Walters

Cardiff University

Daniel R. Weinberger

Lieber Institute for Brain Development, Baltimore, MD, USA
Departments of Psychiatry, Neurology, Neuroscience and Institute of Genetic Medicine, Johns Hopkins School of Medicine, Baltimore, MD, USA

Thomas Werge

The Lundbeck Foundation Initiative for Integrative Psychiatric Research, iPSYCH, Denmark.
Institute of Biological Psychiatry, MHC Sct. Hans, Mental Health Services Copenhagen, Denmark.
Department of Clinical Medicine, University of Copenhagen, Copenhagen, Denmark.
