## Supplementary figures and images for "Combinations of genes at the 16p11.2 and 22q11.2 CNVs contribute to neurobehavioral traits"

### Figure_S2.pdf

#### ASD 22q

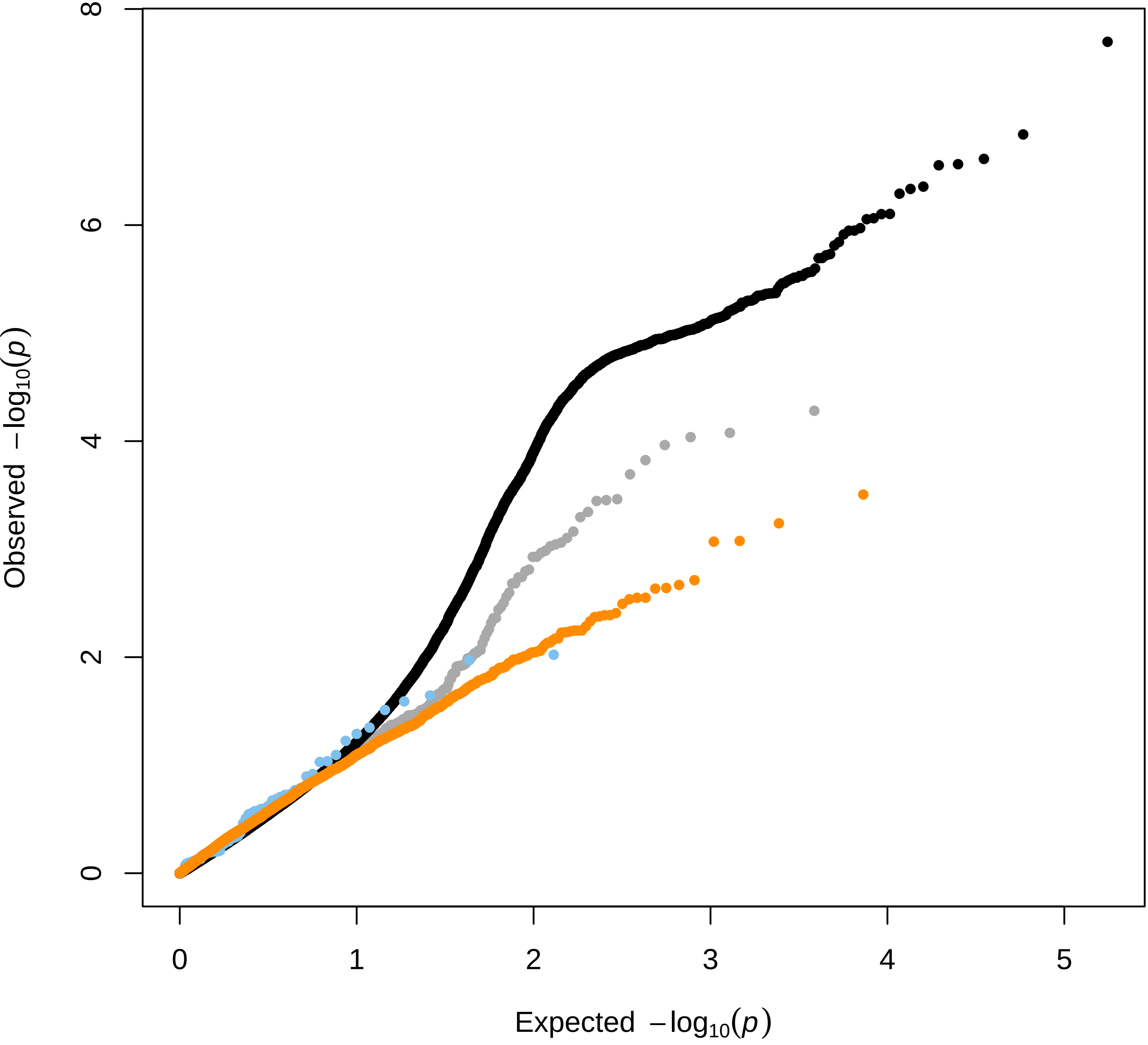

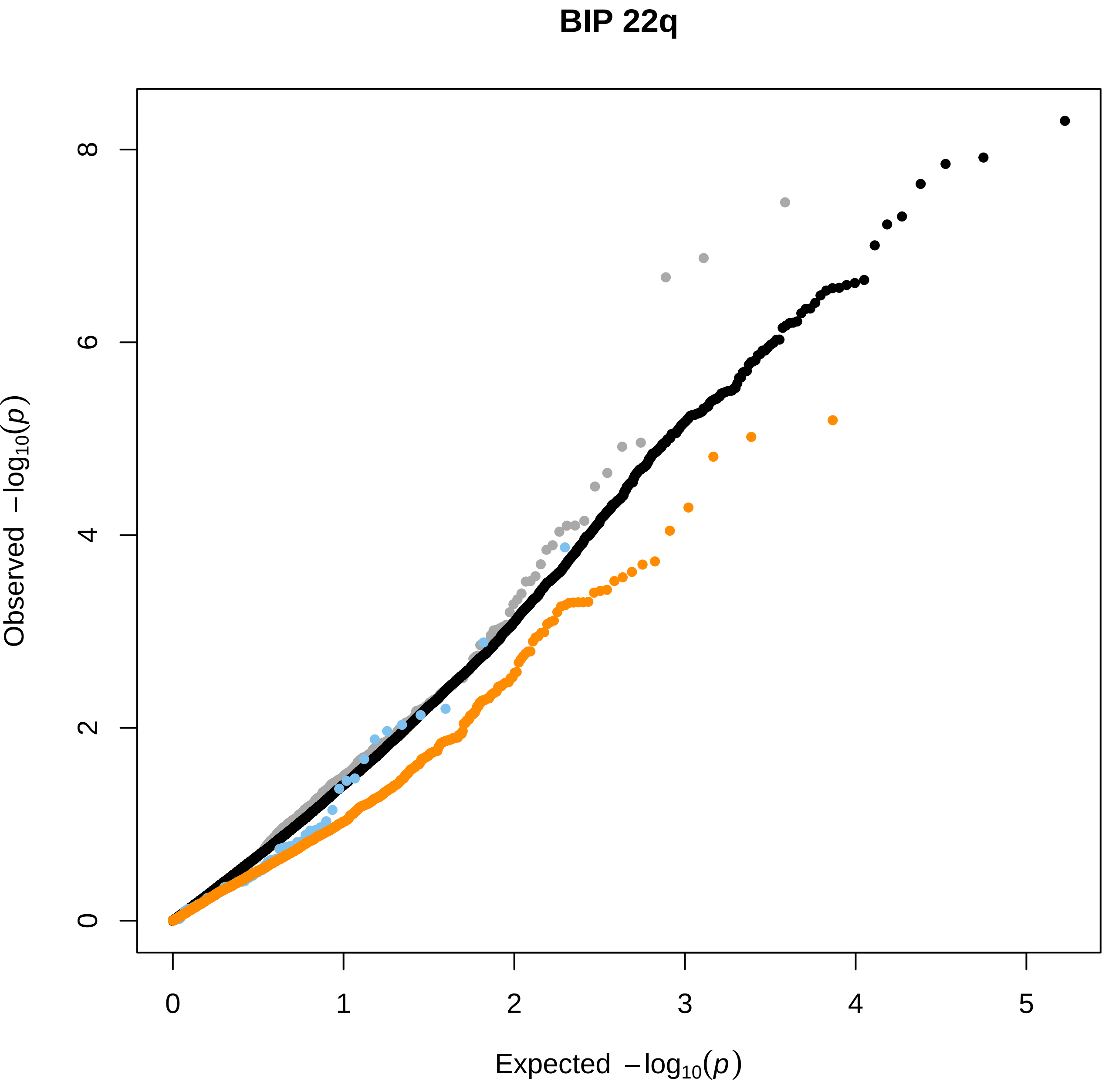

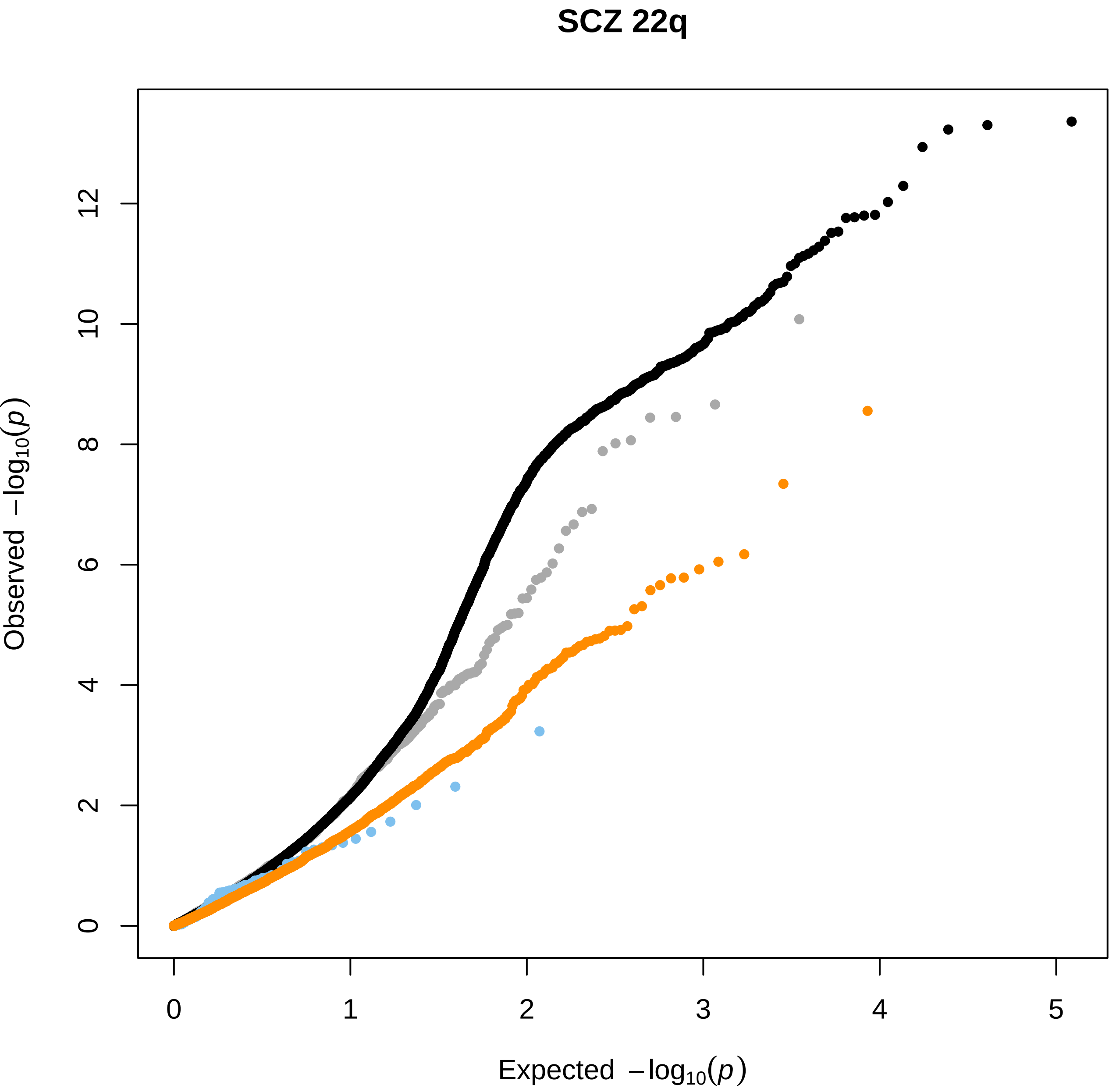

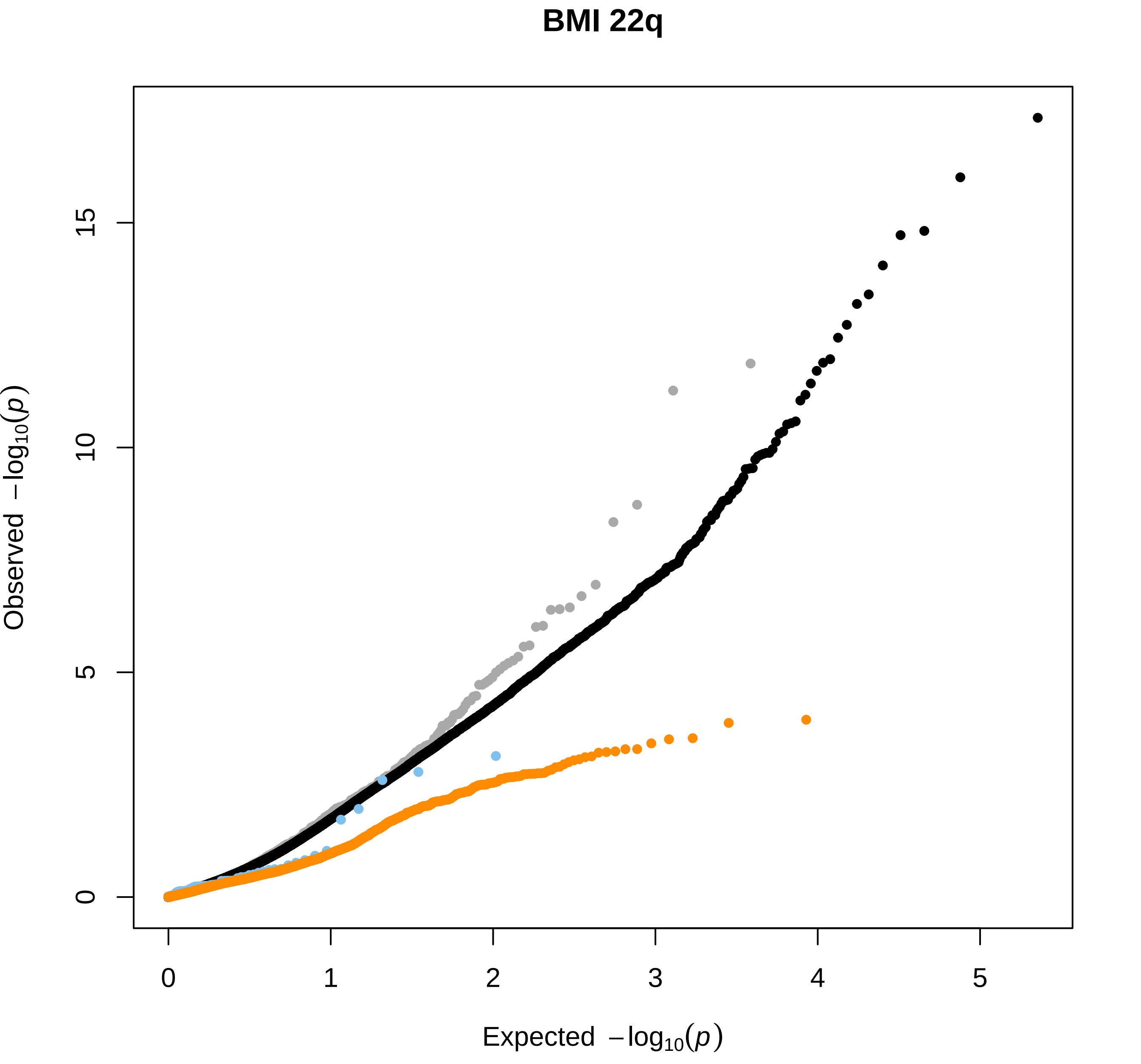

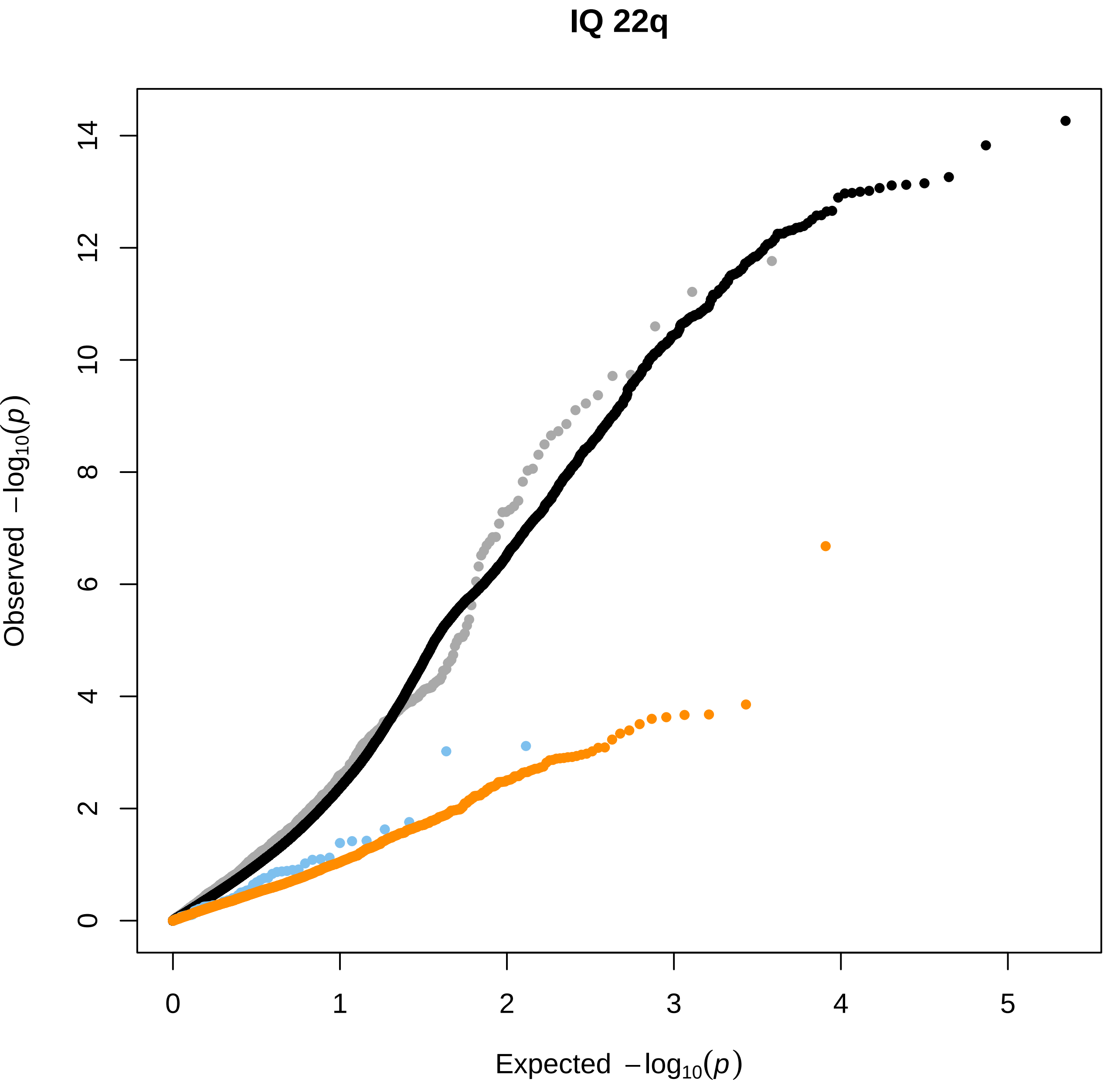

### Figure_S3.pdf

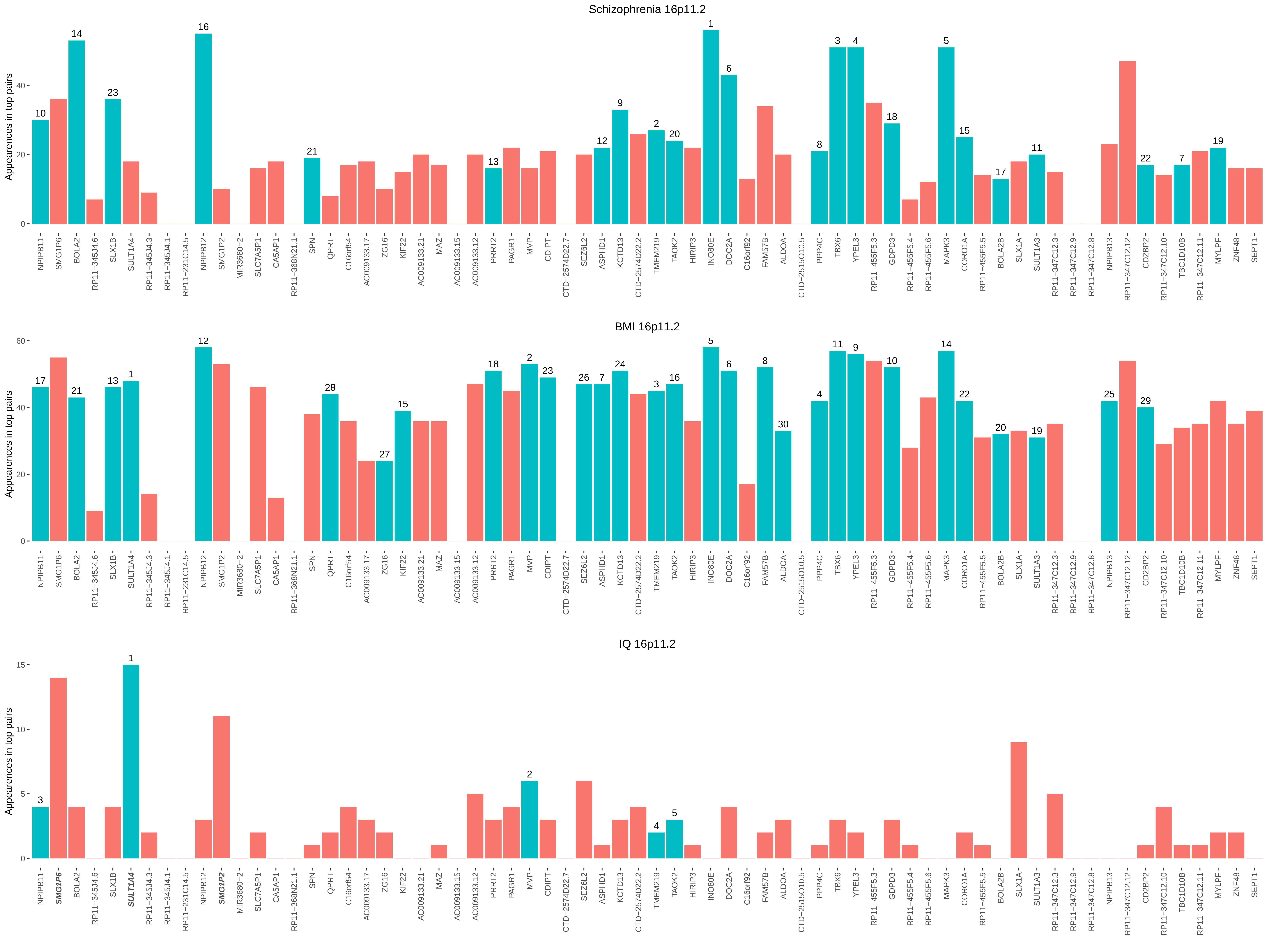

### Figure_S4.pdf

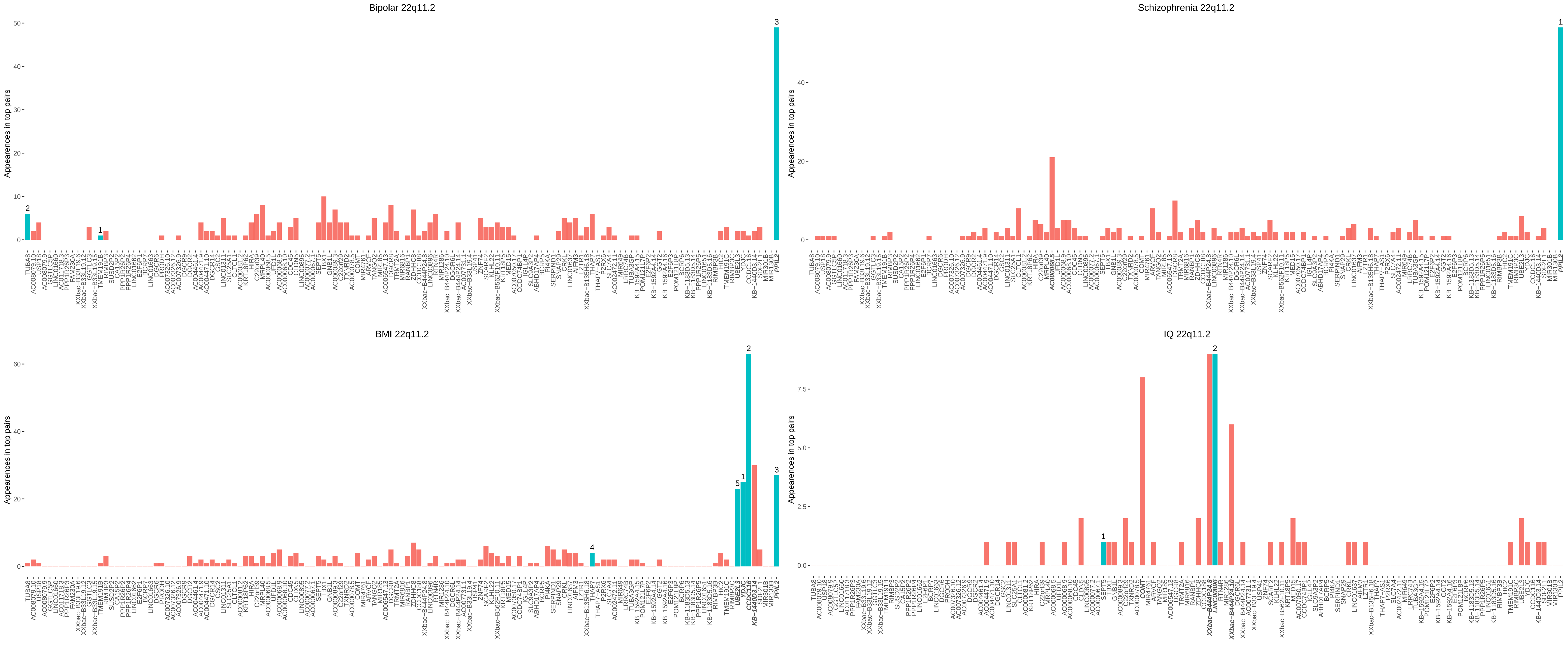

### Figure_S5.pdf

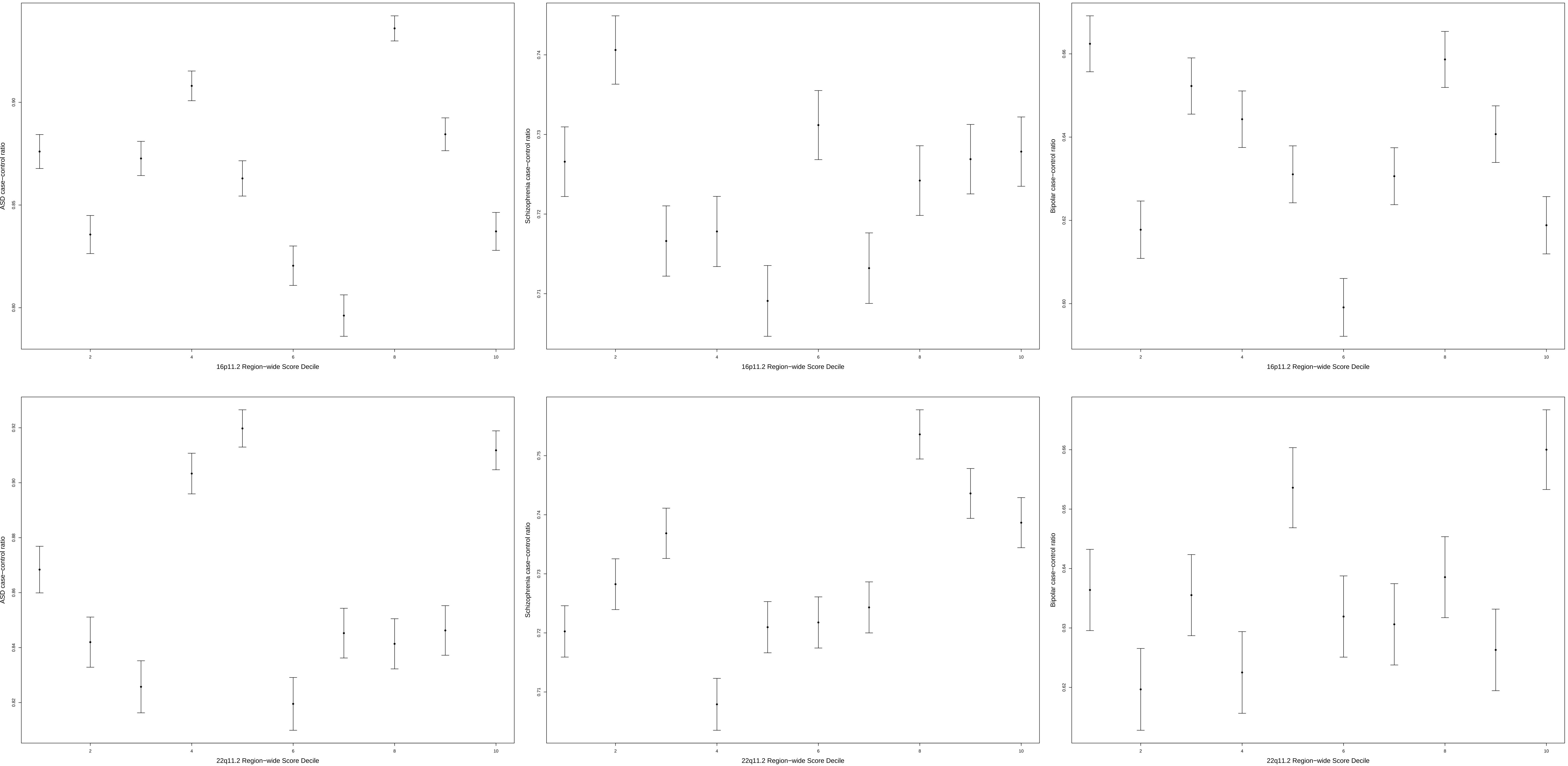
